# AutoSEIR: Accurate Forecasting from Real-time Epidemic Data Using Machine Learning

**DOI:** 10.1101/2020.07.25.20159715

**Authors:** Stefano Giovanni Rizzo, Giovanna Vantini, Mohamad Saad, Sanjay Chawla

**Affiliations:** Qatar Computing Research Institute, Doha, Qatar

## Abstract

Since the SARS-CoV-2 virus outbreak has been recognized as a pandemic on March 11, 2020, several models have been proposed to forecast its evolution following the governments’ interventions. In particular, the need for fine-grained predictions, based on real-time and fluctuating data, has highlighted the limitations of traditional SEIR models and parameter fitting, encouraging the study of new models for greater accuracy. In this paper we propose a novel approach to epidemiological parameter fitting and epidemic forecasting, based on an extended version of the SEIR compartmental model and on an auto-differentiation technique for partially observable ODEs (Ordinary Differential Equations). The results on publicly available data show that the proposed model is able to fit the daily cases curve with greater accuracy, obtaining also a lower forecast error. Furthermore, the forecast accuracy allows to predict the peak with an error margin of less than one week, up to 50 days before the peak happens.

## 1 Introduction

The SARS-CoV-2 virus is a new strain of the Severe Acute Respiratory Syndrome (SARS) species that causes COVID-19. Its long incubation period and the fact that infected individuals are often asymptomatic or exhibit mild symptoms are distinctive features that make spreading the virus relatively easy. The COVID-19 fatality and hospitalization rates are reported to be substantially higher than the regular flu [1] and thus can overwhelm the health services of even advanced developed countries. Thus, to control the spread of the virus, forecasting models for COVID-19 are required for planning and decision making. Despite the online release of daily data from most of the affected countries it has become clear that predicting the future trajectory of the pandemic is a very challenging task [2]. In particular, traditional epidemiological models are less effective when applied to very fine-grain, partial observations, than when applied to historical smooth data.

Many epidemiological parameters are used to model the progress of an epidemic and forecast its future evolution. One important parameter is the basic reproduction number *R*_0_, which defines the average number of new infections resulting from one individual carrying the virus in a completely susceptible population. It has been shown that the measured *R*_0_ as well as other epidemiological parameters vary widely across different countries and studies. As a result, current estimation of *R*_0_ for COVID-19 epidemic ranges from 1.4 to 6.49[3]. Moreover, the fact that the countries have responded in different manners to prevent its spread makes it difficult to infer the effective reproduction number and other transmission parameters either before or after intervention. One of the limits of traditional compartmental models (e.g., SEIR) is the assumption that the epidemic parameters do not change over time. In practice, most countries have imposed lockdowns and restrictions to mobility, together with physical distancing guidelines, which affected the average number of contacts and the transmissibility of the virus. Models that do not consider the change in transmissibility resulting from lockdowns have not produced accurate predictions [2, 4].

Once the transmission parameters are accurately estimated, SEIR models can be applied to simulate the evolution of the epidemic. One key challenge in this process is simulating the SEIR compartments while the only available data are the newly reported cases and standard compartments are not observable. Since the newly reported cases are only a fraction of the real infected individuals, in some studies the population has been artificially reduced to a much lower value in order to obtain reasonable results from an SIR model [5]. Moreover, confirmed cases are usually reported a long time after the infection occurs, which explains why the effect of intervention (reduction of new infections) is only observable after at least two weeks. Unless we choose an incorrect incubation period of around two weeks [6], which is much longer than the estimated median of five days [7], it is not possible to fit the observed data with a standard SEIR model. A time gap can be manually fixed [5, 8], for example shifting the prediction or the original data, although an ideal solution would consider this gap to be country-dependent and it should learn the gap length automatically from the data. In general, traditional models need to be extended to cope with all the above challenges.

In this paper, we propose a new extended SEIR model with machine learning parameter fitting, we call AutoSEIR. The main contributions of AutoSEIR are: (i) we propose a new compartment-based model to capture the unique characteristic of COVID-19. In particular we extend the classical SEIR model by introducing two additional compartments, Tested and Untested. The Untested compartment is not observable as the proportion of the population that is being tested continues to be relatively small; (ii) we introduce a testing delay parameter and a confirmed compartment to capture the reporting delay, which usually occurs between the infection and the reporting stage of each case; (iii) we consider two different transmissibility values, before and after intervention, while fitting the whole time series, conversely to other methods that fit each period separately, obtaining a discontinuous result [9]; and finally (iv) we extend a recently introduced proximal optimization framework for parameter inference for ODEs, using automatic differentiation, to account for interventions and state variables that are not observable. In this study, we evaluate the method for accuracy in parameter fitting, forecast capabilities, peak prediction, and robustness to outliers in the data. The results and daily-updated forecasts are publicly available on our website (https://covid-research.qcri.org/seir/).

## 2 Material and Methods

### 2.1 Proposed Extension of SEIR Model: AutoSEIR

Our proposed extended SEIR model contains seven compartments: S (Susceptible), E (Exposed), I (Infected), T (Tested), U (Untested), C (Confirmed) and R (Removed) (Figure 1). Our model accounts for the delay between symptom onset, getting an RT-PCR test, and also the outcome of the RT-PCR test.

**Figure 1:**
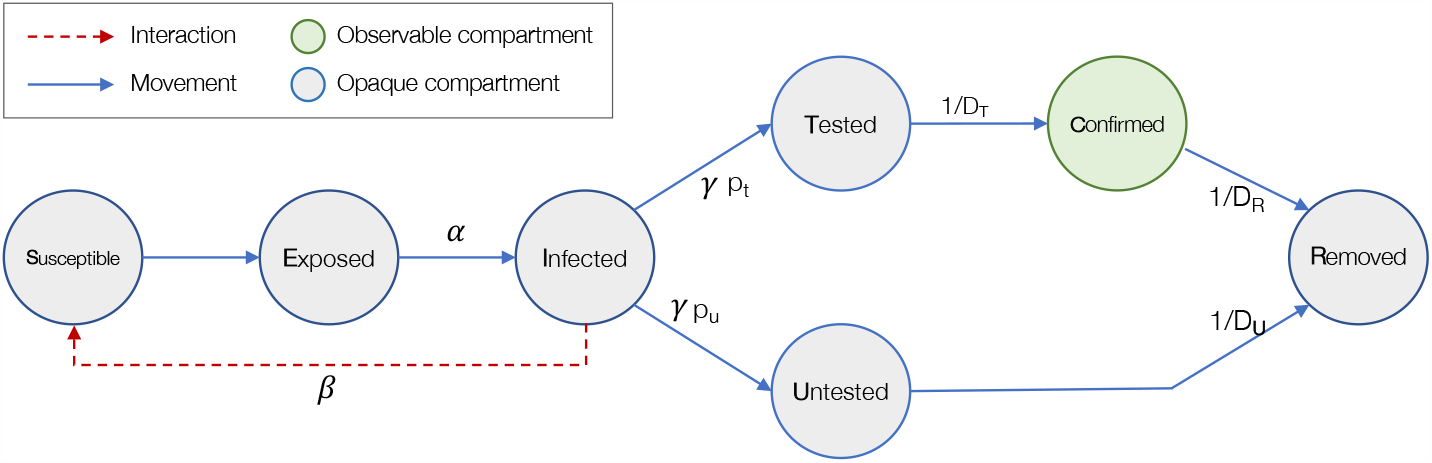
Diagram of AutoSEIR. Only the confirmed compartment is observed.

The dynamics of the compartments are defined by the following ordinary differential equations (ODEs):

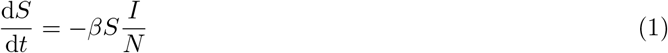

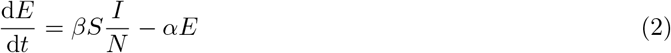

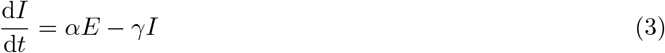

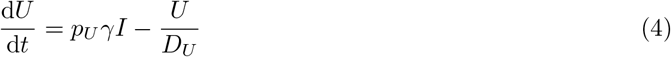

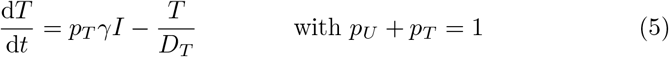

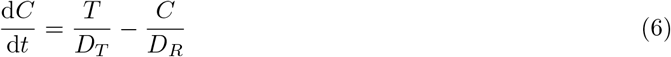

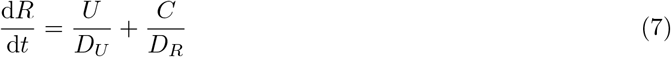

The model contains several parameters, fully described in the following section. In the above equations and in Figure 1 we can separate our parameters in three categories: (1) the transmissibility *β* that controls the rate of susceptible individuals getting infected after getting in contact with an infected individual, (2) the time rate parameters {*α, γ, D*_*T*_, *D*_*R*_, *D*_*U*_} that control the timing of moving from a compartment to the next, and (3) the ratios *p*_*T*_ and *p*_*U*_ to split between tested and untested compartments. Since we have two different rates before and after the social distancing is enforced, the *β* parameter is the only parameter that varies with time.

### 2.2 Parameter Fitting

A distinct feature of our approach is that we can estimate the parameters even when several ODE equations are un-observable. In practice, we will assume that only the compartment of confirmed cases is observable, which happens after individuals have been tested.

Given the defined ODE system, the complete state over the seven compartments at time *t* is *X*(*t*) = *x*_*S*_(*t*), *x*_*E*_(*t*), *x*_*I*_ (*t*), *x*_*U*_ (*t*), *x*_*T*_ (*t*), *x*_*C*_(*t*), *x*_*R*_(*t*). The state depends on two sets of parameters. One set has fixed values, and one set must be estimated. The vector *θ* is the vector of free parameters that we infer through optimization and it includes the following:

1. *I*_0_: the initial infected, i.e., *x*_*I*_ (0) = *I*_0_.
2. *R*_0_: basic reproduction number, assuming a completely susceptible population.
3. *R*_*t*_: basic reproduction number resulting from social distancing, also assuming a completely susceptible population.
4. *D*_*T*_ : delay of testing, i.e., time from the end of infectious period to the time the case is officially reported.
5. *p*_*T*_ : probability of an infected case to be tested. This is equal to 1 − *p*_*U*_, where *p*_*U*_ is the probability of unreported cases.

The fixed parameters are:

1. *D*_*U*_ : recovery duration for the unreported cases, from the end of the infectious period to the removed/recovered compartment.
2. *D*_*R*_: recovery duration for the reported cases, from the reporting of the case to the removed/recovered compartment.
3. 1*/α*: incubation period.
4. 1*/γ*: infectious period.
5. *t*_*s*_: timestep when social distancing measures becomes effective.

The rationale of fixing the above parameters is that the time of social distancing is a known datum, while the timing of recovery compartment was not crucial to the scope of this work and can be more easily estimated from reported data. In a scenario where the estimation of recovery time is a key factor, such as in policy-making for hospital capacity management, *x*_*R*_ should be also observable and *D*_*U*_, *D*_*R*_ can be included in the parameter vector *θ*.

The initial timestep *t*_0_ is variable, given by *t*_*obs*_ −(*α* + *γ* + *D*_*T*_), thus it depends on the current parameter values.

#### ODE Solver

In order to obtain the number of cases, given the defined system of ODEs and the current parameters vector *θ*, we solve numerically the ODEs using the midpoint method. The solver takes as input the initial state X(0), the system of ODEs *f*_*θ*_(*X*_*t*_, *t*), and iterates over *t*, approximating each *X*(*t* + 1) using the midpoint rule:

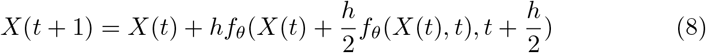

The initial *X*(0) = {*x*_*S*_(0), *x*_*E*_(0), *x*_*I*_ (0), *x*_*U*_ (0), *x*_*T*_ (0), *x*_*C*_(0), *x*_*R*_(0)} is set as {*N* − *I*_0_, 0, *I*_0_, 0, 0, 0, 0}, where *N* is the total population of the considered country or area, and *I*_0_ ∈ *θ*, the initial number of infectious subjects, is part of the parameters vector that will be fitted.

#### Loss function

A loss function is defined to measure the error of the simulated results with respect to the real acquired data. In the real data, not all the compartments of *X* are observable, instead only the ground truth for the new reported cases is always available, which we call *y*(*t*). Note that the corresponding variable in our system is not directly available, as we simulate the state of currently infected, but we do not keep track of the newly confirmed cases at time *t*. This variable can be defined as:

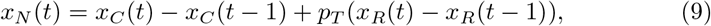

that is the new cases are given by the current number of confirmed cases *x*_*C*_(*t*), removing the previous step confirmed cases *x*_*C*_(*t* − 1), and adding the cases that left the confirmed department, *p*_*T*_ (*x*_*R*_(*t*) −*x*_*R*_(*t* −1)).

The loss function over all the states is

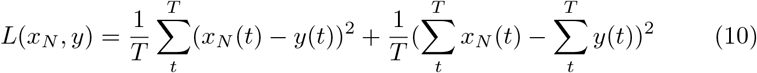

where *y*(*t*) is the observed confirmed cases at discretized timestep *t* and T is the entire dataset period. In this function we are considering the point-wise error of prediction as well as the cumulative error so that we optimize for both the daily cases and the total cumulative cases.

The loss surface as a function of the parameters pair {*R*_*t*_, *p*_*T*_} on the Italian dataset is shown in Figure 3. In this dataset the number of new cases started to decrease when 0.1% of the population was confirmed as infected, as a result of the imposed restrictions. This decreasing curve can only be fitted either by having an *R*_*t*_ smaller than 1, or by considering that in reality a high percentage of the population has been infected but not tested, that is having a very small value for *p*_*T*_. This is reflected in the surface plot where the fitting loss increase exponentially for *R*_*t*_ *>* 1 and high *p*_*T*_. The lowest loss is instead obtained when *R*_*t*_ is around 0.8 and the probability of testing is around 0.2.

**Figure 2:**
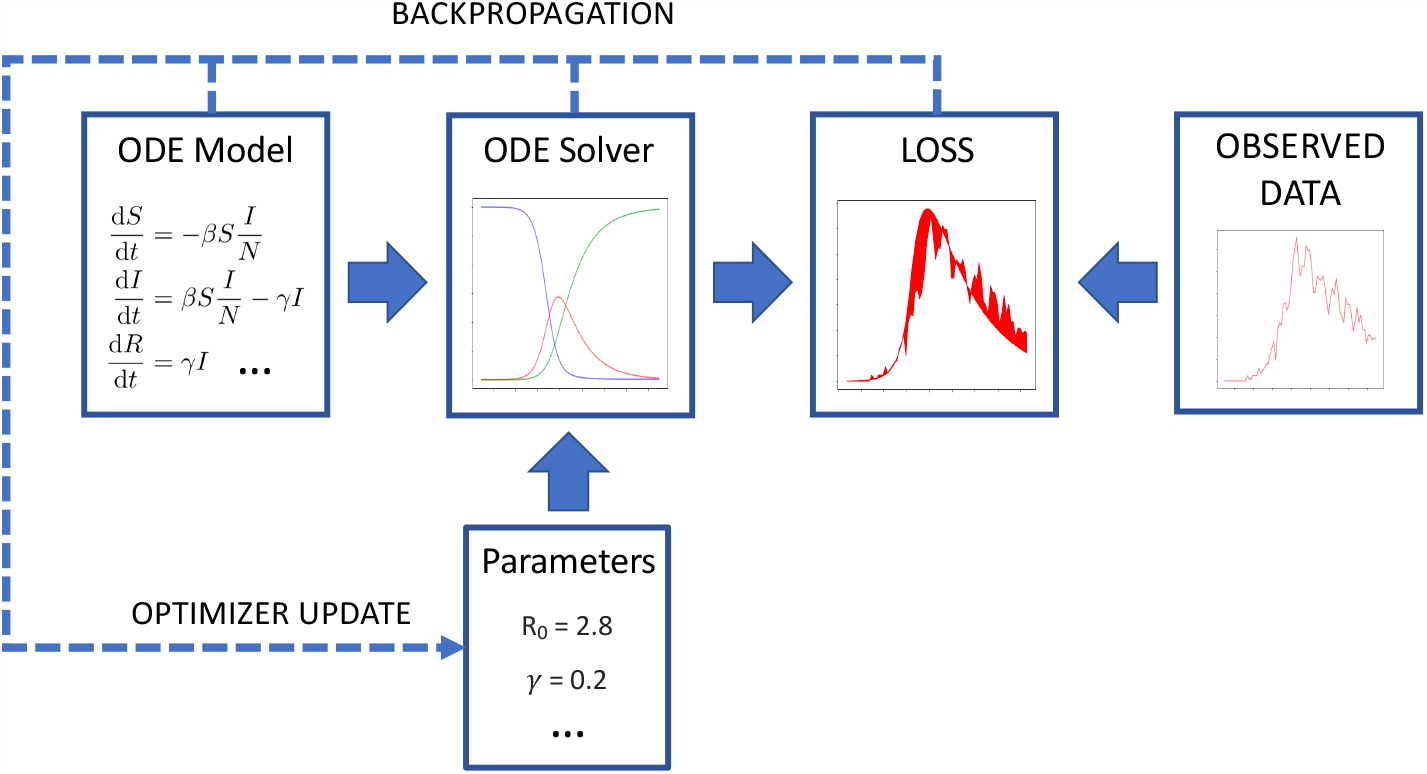
Diagram of the full approach for parameter fitting using backpropagation.

**Figure 3:**
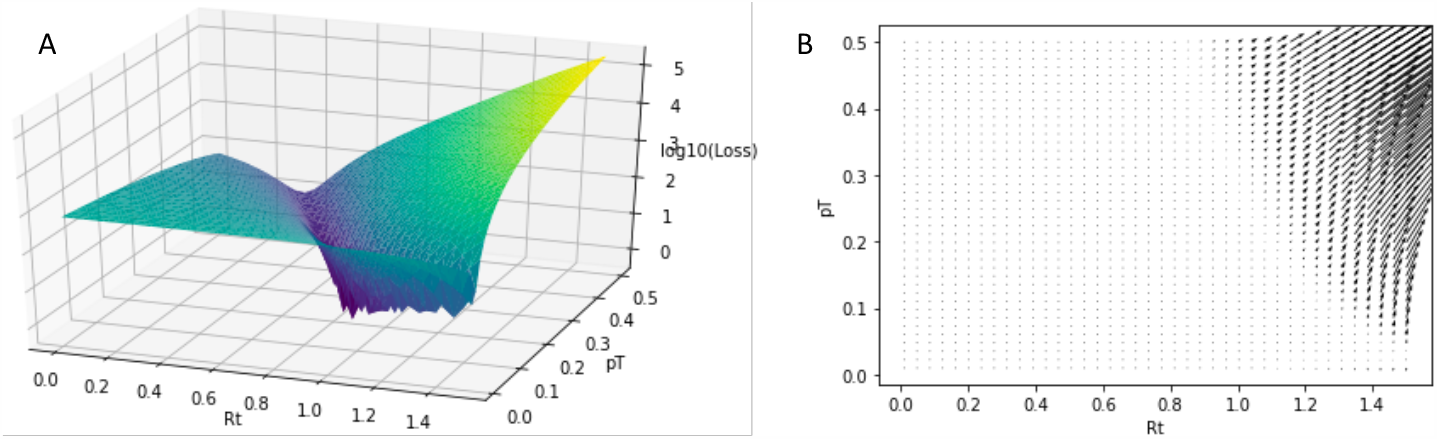
Loss surface (A) and loss gradients (B) with respect to the parameters {*R*_*t*_, *pT*}. Data of Italy daily cases is used to measure the loss.

#### Constrained optimization

The optimization problem is defined by the constrained minimization objective

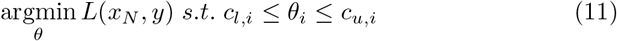

where *c*_*l,i*_ and *c*_*u,i*_ are lower and upper bounds enforced on the parameter *θ*_*i*_. To compute the gradient of the objective function with respect to the model parameters, we use the automatic differentiation python package `autograd`.

#### Timing

A unique feature of the proposed method is that, in the ODE solver, the simulation starts a period of time before the first observed cases. This is required in order to match the confirmed compartment’s count to the confirmed cases in the observed data, given that the defined ODE implies a sequence of movements that starts from the susceptible compartment and reaches the confirmed compartment after a period of time. The average duration of this time period is given by the sum of the average periods between the infection compartment and the confirmed compartment, given by the latent/incubation period, the infectious period, and the test delay, which is 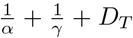. Note that, contrary to other approach where a time gap is manually fixed [8], in our approach the duration is automatically set based on the fitting of *D*_*T*_. The time discrepancy between the ODE and the observed data is illustrated in Figure 4. In this chart, which represents the Hubei province initial cases, the first observed data point in the dataset is on January 22. Nevertheless, to fit such data, the infection simulation of the ODE solver starts on January 10 (i.e., 12 days before). This time shift accommodates the period of time needed to go from the first, yet unobserved cases, to the 400+ cases first observed after almost two weeks.

**Figure 4:**
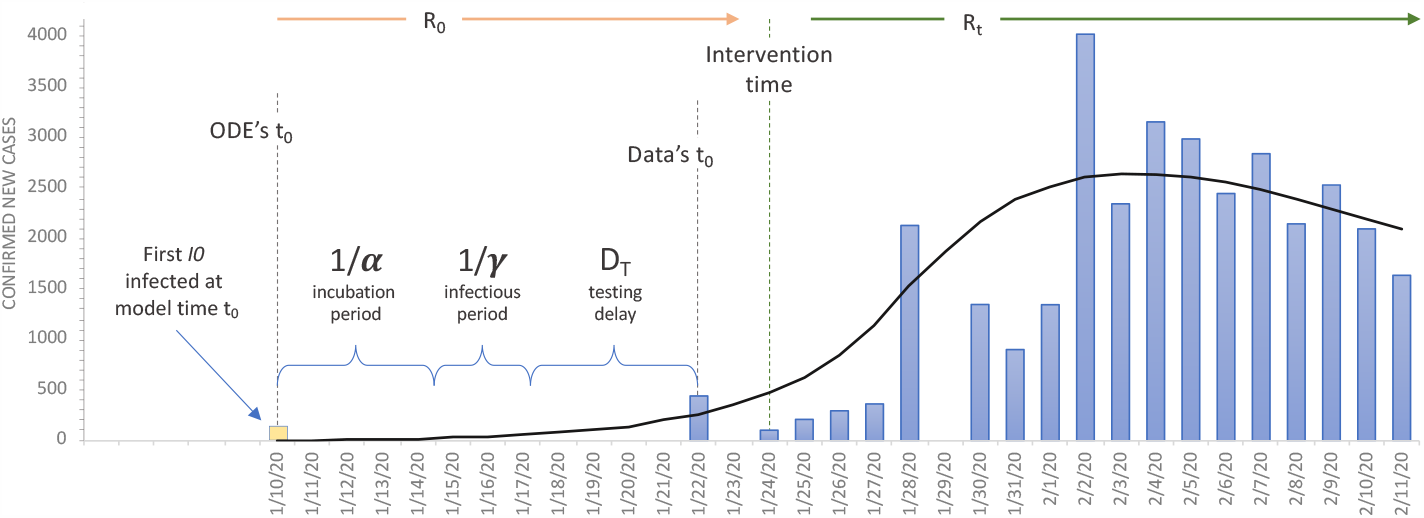
Timings of ODE model and observed data. The ODE’s first timestep *t*_0_ depends on the given observed data’s *t*_0_ and the durations of incubation period, infectious period and testing delay. Time periods and initial infected *I*_0_ are all optimized parameters.

### 2.3 Experimental data and setup

The data used in our experiments are collected from the three sources shown in Table 1 with the corresponding areas of interest. Lockdown intervention dates have been extracted from newspaper sources of each country, summarized on the related Wikipedia page of COVID-19 pandemic lockdowns (https://en.wikipedia.org/wiki/COVID-19pandemiclockdowns).

**Table 1:** Data source repositories used for experiments, by area.

| Data Source | Area | Repository URL |
| --- | --- | --- |
| New York Times | New York state | https://github.com/nytimes/covid-19-data |
| Italy Civil Protection | Italy | https://github.com/pcm-dpc/COVID-19 |
| Johns Hopkins | Hubei | https://github.com/CSSEGISandData/COVID-19 |
|  | S. Korea |  |
|  | Spain |  |
|  | Germany |  |
|  | France |  |
|  | India |  |
|  | U.K. |  |

For all the experiments, we setup the same upper and lower bounds constraints and initial values (Table 2). The constraints are enforced in order to avoid negative values as well as unrealistic high values. Initial values are reasonable values to start the numerical optimization. Both initial values and constraint ranges are shown in Table 2. All values are constant except for the initial number of infected (i.e., the observed cases at the first time step *y*(0)).

**Table 2:**
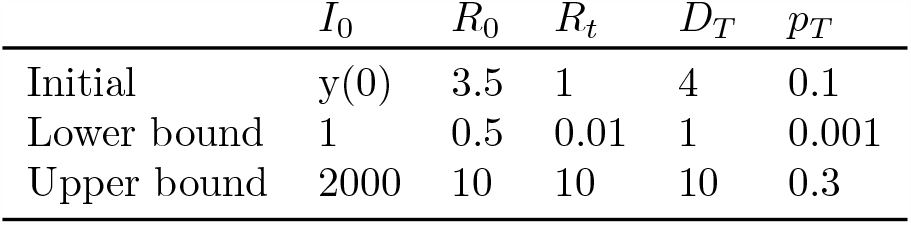
Initial values, lower bound and upper bound constraint for all the free parameters of the optimization.

## 3 Results

### 3.1 Ablation Study

We perform ablation experiments to investigate the impact of each proposed extension of the SEIR model that makes up our complete approach. Specifically, we evaluate the accuracy of the model under the following five configurations:

1. **Full model**. This is the full model presented in Section 2.
2. **No** *p*_*T*_. In this model we remove the probability of testing, assuming that all the infected cases are tested and observable.
3. **No** *D*_*T*_. This model has no test delay, thus all the infected cases are reported as soon as they are tested.
4. **No** *R*_*t*_, *D*_*T*_. In this model the intervention does not have any effect, thus all parameters learned for data points before and after intervention are the same. Additionally, the model has no test delay feature, meaning that all the infected cases are reported as soon as they are tested.
5. **No** *p*_*T*_, *D*_*T*_. In this model both the probability of testing and the test delay are removed. All infected cases are tested, and reported without any delay.

We evaluate the described models in two different experiments:

- **Fitting**. In this experiment we evaluate the ability of the models of fitting the observed data. The complete data up to a recent date is provided for training, and the fitting of the training data is evaluated. The purpose of this experiment is to show that without any of the missing feature the model would not be capable of fitting real data, with the exception of fitting parameters with non-realistic values.
- **Forecast**. We design this experiment to evaluate the predicting capability of the models. We train the models with data spanning from the start of the epidemic until one to six weeks after the intervention date. For testing, we consider the week following the end of the training data, and we measure the prediction accuracy of each model on this unseen 7-points time series.

The above experiments are carried out on the datasets of U.K., Italy, U.S., and New York. Results of the ablation and forecast experiments are shown in Figure 5 and 6, respectively. Fitting results of the full model are shown in the first row. For each fitting, the root mean square error (RMSE) is given for the whole time series. In all the experiments except one, the result of the full model has the lowest error among the models without one or more extensions. Only the ’No *D*_*T*_ ’ model for the Italy dataset has a slightly lower error. However, the inferred rate of testing parameter was unrealistic (0.7%), for a country where the testing ratio has been estimated to be in the range of 10%-33% [10, 11]. The same ’No *D*_*T*_ ’ model yields a good fitting for the rest of the datasets as well, with the downside of estimating an *R*_0_ parameter to be higher than the known value. The rest of the models were not capable of reasonably fitting all the datasets. In particular, the UK dataset could not be fitted without using the testing ratio *p*_*T*_ or the after-intervention *R*_*t*_ parameter. For the U.S. dataset, only the model ’No *R*_*t*_, *D*_*T*_ ’ failed to fit the observed data by a large margin.

**Figure 5:**
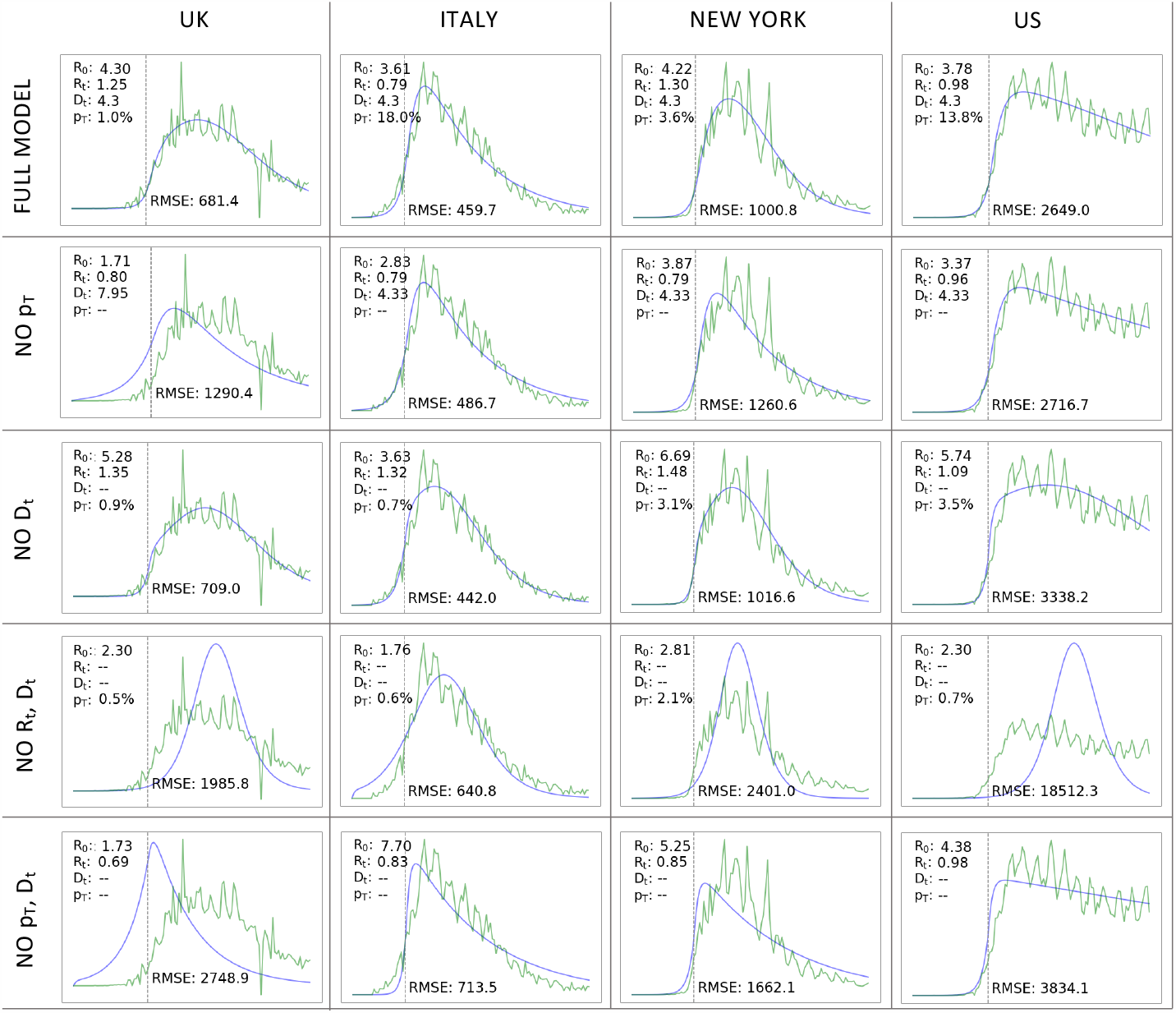
Results of ablation experiments on fitting for UK, Italy, New York and U.S. All the features combined (full model) produce the best fitting while using more reasonable parameters.

**Figure 6:**
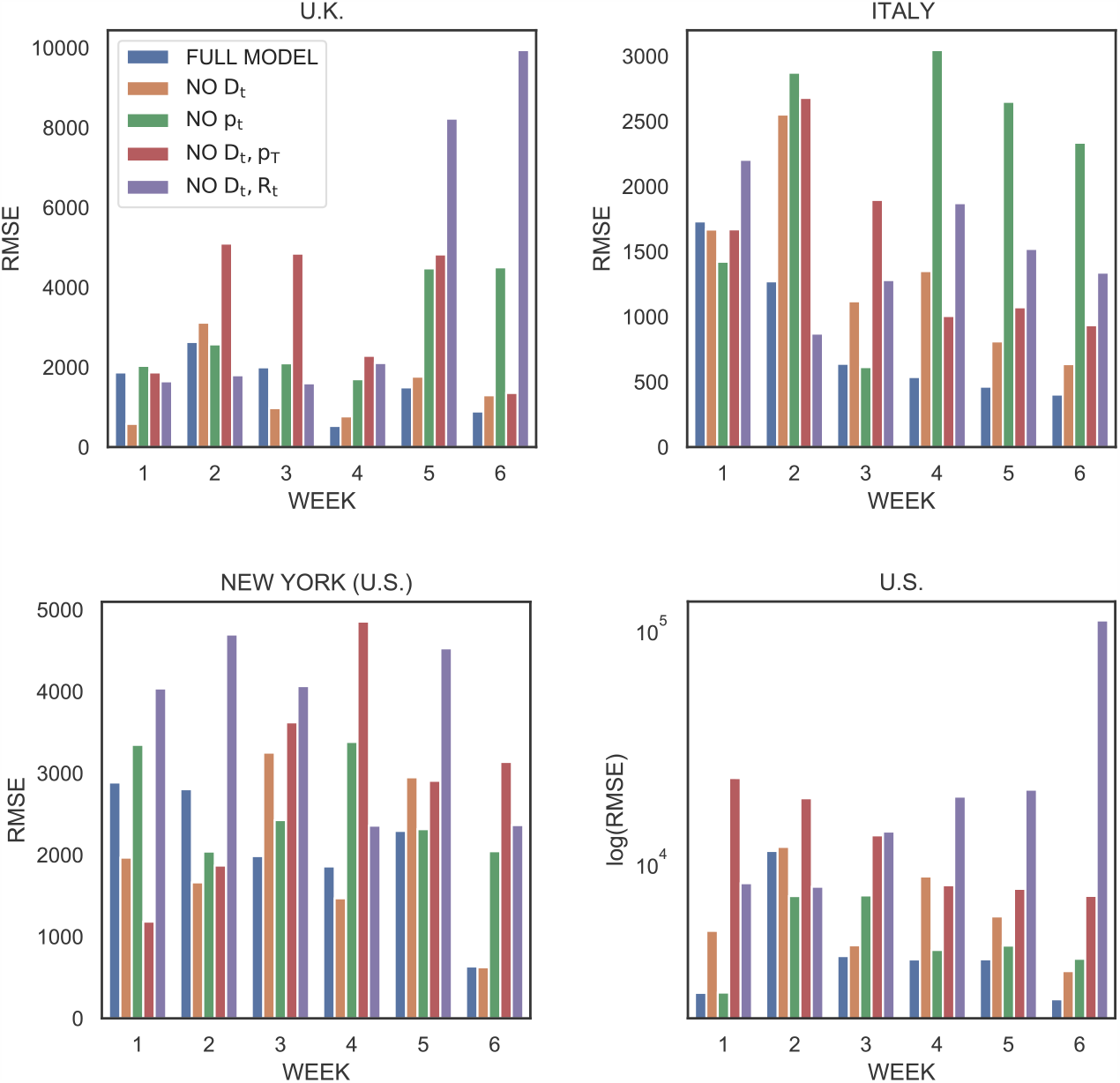
Error on next week forecast for Italy, Germany, and UK datasets.

For the forecast experiment, we show the RMSE of the five models for each period used as training set (week from 1 to 6). The full model result, represented with a solid blue line, exhibits the lowest error in most of the configurations. The performance of the remaining models vary widely across different countries datasets and periods of time without any particular pattern. Logically, our results show that the full model has a decreasing trend in error with respect to the increase of the training data, showing that the more historical data is available to the model, the better it predicts future points. This suggests that the full model is good at generalizing the epidemic dynamic, while the remaining models are overfitting the observed data in an unpredictable manner, lacking a robust underlying model.

### 3.2 Robustness

The availability of almost real-time data for the ongoing COVID-19 epidemic has resulted in the occurrence of several fluctuations, outliers and anomalies in the public datasets, which represents an issue for parameter fitting among other challenges. One example of these anomalies is the 600% surge in cases observed on February 12 for the contagion data in Hubei province, where 13,332 cases were observed in one day. The anomaly was caused by a policy change on the counting of cases, when the co-presence of symptoms and chest infection from CT scan was reported as confirmed case. To test the robustness of our model to these contexts, we train on the Hubei province data until February 18, thus including the spike, and we generate the forecast of 6 weeks to validate both fitting and prediction accuracy. In Figure 7, the results are shown together with the overlapping observed data represented with a red solid line. The figure shows that the fitting of the observed data is not misled by the outlier, moreover the resulting forecast has been reasonably accurate.

**Figure 7:**
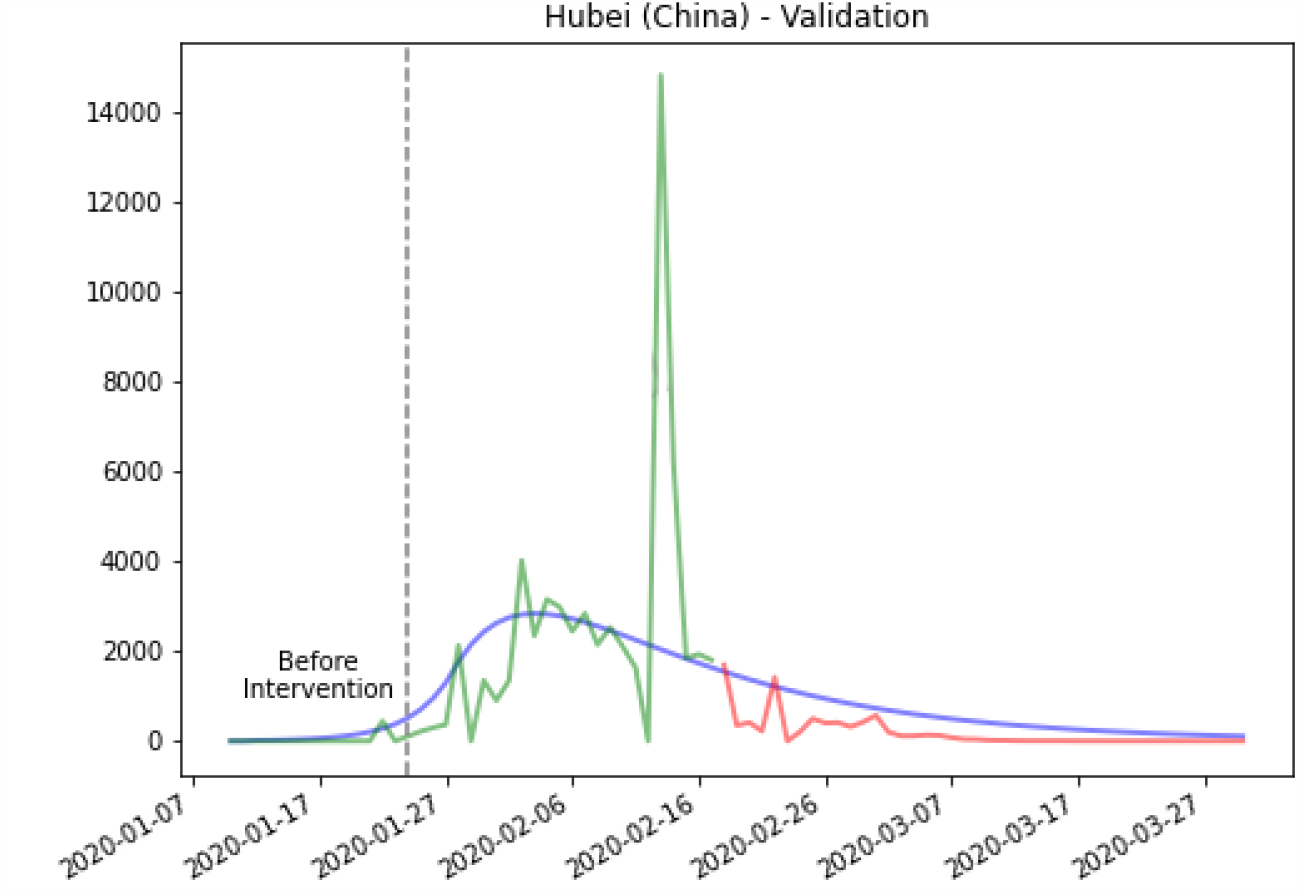
Hubei validation. Training data up to February 18. Note: despite the peak on February 12, resulted from a change in the confirmation policy, the model show robustness to outliers.

### 3.3 Peak prediction

In this experiment we evaluate the ability to predict the date when the peak occurs. Given the predicted new cases for each time step, *x*_*N*_ (*t*), we obtain the predicted peak timestep as

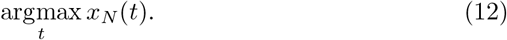

We select the COVID-19 new cases dataset of Qatar as a case study. We run the parameter fitting using as training data starting from 2 months before the peak up to the day before the peak, and we measure the difference with the actual peak date. This difference is defined as

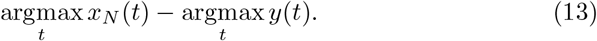

In Figure 8 we show the result for each day. For example, 35 days before the real peak happened (-35 on the figure) the method was able to predict with exact accuracy the peak date. The accuracy is high on average, with a mean absolute error of 10 days. The model has been fooled by a false peak around 30 days before the real peak, however it managed to well re-tune its prediction after more days of data has become available.

**Figure 8:**
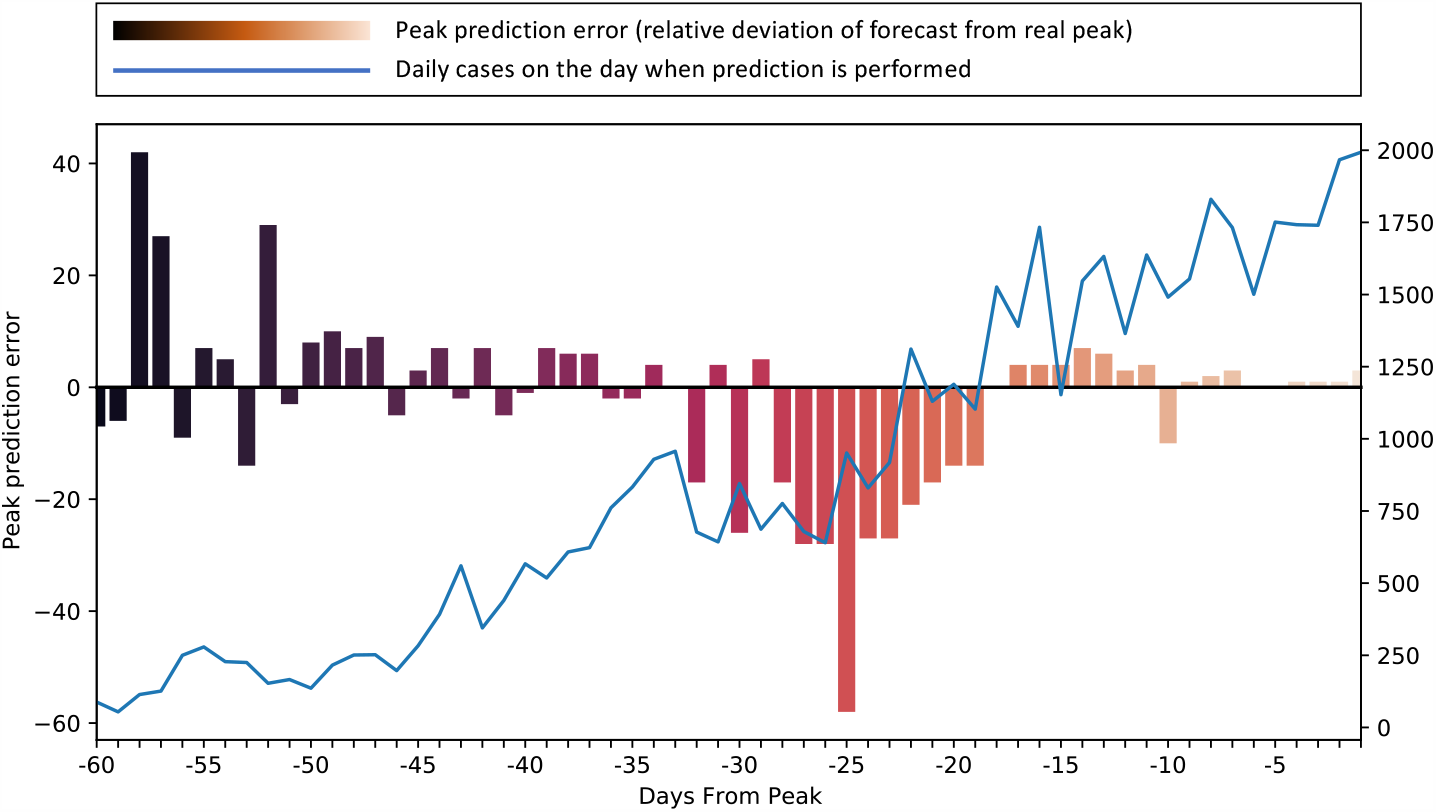
Peak prediction on Qatar cases. Starting from 60 days before the peak, we forecast the peak date every day. Relative error on forecast is provided as a bar for each day. Solid line represents the daily new cases. The prediction has high accuracy except when close to a deceitful peak, which happened around-30 days with respect to the real peak.

## 4 Conclusions and Discussion

We have proposed a novel approach that combines an extended compartmental model, to address the unique features of COVID-19 pandemic and of the available data, with a novel machine learning technique for parameter fitting on ODE systems with partial observations. This approach allowed us to obtain a state-of-the-art forecast accuracy and peak prediction. The resulting forecasts have been used in government settings and international media outlets as a trusted source for the outbreak projections. Machine learning has been successfully applied to epidemic curve prediction in the past, with methods that treat the problem as a standard supervised time series forecasting task [12] and a wide literature on estimating the epidemic parameters for predicting the future of an ongoing outbreak, especially for influenza [13]. The COVID-19 emergence, together with the readiness and availability of related data, has attracted great attention on epidemic modeling research, in the pursuit of an accurate method to predict the future outcome of the pandemic. Several approaches have been proposed in the recent literature to fit the confirmed cases observed in public datasets. In general, it has been shown that epidemic forecasting and peak prediction is a very challenging task that leads to very inaccurate results with traditional SEIR optimization approaches, especially when interventions are involved [2, 4].

In [14], an architecture composed of a neural network and a time integrator is trained to to fit infected and deaths data in a traditional SIRD model, however this is not used to produce forecasts. Also a full library has been presented, named PyRoss [15], to model the epidemic and fitting the data using Bayesian parameter inference and model selection. To cope with the artificial factors that influence the daily data, the SUQC model (Susceptible, Un-quarantined, infected, Confirmed infected) has been proposed instead of SEIR [16], with a loss function optimized using the interior-point method. In [9], an extension of SEIR is proposed, to additionally model unreported infections and hospitalized subjects. The observed time series is split into several period, to account for the change given by restrictions, and the parameters of each subset are fitted in isolation. This results in a discontinuous fitting and inaccurate long-term forecast than using a unified contiguous approach as we propose. More complex extensions to SEIR for COVID-19 include the SEIQRDC model [17], where the exposed population is able to infect during the latent period, confirmed cases are quarantined and susceptible are partially confined, and a compartmental model that represents mild, severe and critical infections separately, as well as severe and critical disease [18].

Other than SEIR models, more simple models combining power law and exponential law have been used for fitting, which obtain more accurate results than non-extended compartmental models for forecasting [19]. Monte Carlo simulations have been also applied to model the cumulative cases and predict the reduction of fatalities [20].

The problem of the unconfirmed cases and the time lag in case confirmation has been addressed in other works by reducing the initial population to a small fraction and shifting the date of the first cases [5, 8]. With the proposed approach, both the real and confirmed cases are simulated, while only the confirmed cases are used to compute the fitting error.

Automatic differentiation of ODE solvers has been used for parameter fittings [21], however, conversely to our method, it is assumed that the values of all the variables in the ODEs, for all the time steps, are available. In the problem we address, only one of the variable can be observed, this required a novel solution.

As a future direction, more compartments can be included in the SEIR model (e.g., Hospitalized, Vaccinated). Other important demographic factors can be also integrated in the model such as age, sex, mobility data flows, between areas in a city and regions in a country, to model the effect of urban mobility and national transportation lockdowns, with the final goal of supporting the decision making surrounding the enforcement and the lifting of restrictions.

## Data Availability

Datasets are publicly available.

## References

[1] R. E. Jordan, P. Adab, and K. Cheng, “Covid-19: risk factors for severe disease and death,” 2020.

[2] W. C. Roda, M. B. Varughese, D. Han, and M. Y. Li, “Why is it difficult to accurately predict the covid-19 epidemic?,” Infectious Disease Modelling, 2020.

[3] Y. Liu, A. A. Gayle, A. Wilder-Smith, and J. Rocklöv, “The reproductive number of COVID-19 is higher compared to SARS coronavirus,” Journal of Travel Medicine, vol. 27, 02 2020.

[4] T. Kuniya, “Prediction of the epidemic peak of coronavirus disease in japan, 2020,” Journal of clinical medicine, vol. 9, no. 3, p. 789, 2020.

[5] D. Fanelli and F. Piazza, “Analysis and forecast of covid-19 spreading in china, italy and france,” Chaos, Solitons & Fractals, vol. 134, p. 109761, 2020.

[6] C. MacIntyre, “On a knife’s edge of a covid-19 pandemic: is containment still possible?,” Public Health Research Practice, vol. 30, 03 2020.

[7] A. R. Akhmetzhanov, K. Mizumoto, S.-m. Jung, N. M. Linton, R. Omori, and H. Nishiura, “Epidemiological characteristics of novel coronavirus infection: A statistical analysis of publicly available case data,” medRxiv, 2020.

[8] Z. Yang, Z. Zeng, K. Wang, S.-S. Wong, W. Liang, M. Zanin, P. Liu, X. Cao, Z. Gao, Z. Mai, J. Liang, X. Liu, S. Li, Y. Li, F. Ye, W. Guan, Y. Yang, F. Li, S. Luo, Y. Xie, B. Liu, Z. Wang, S. Zhang, Y. Wang, N. Zhong, and J. He, “Modified seir and ai prediction of the epidemics trend of covid-19 in china under public health interventions,” Journal of Thoracic Disease, vol. 12, no. 3, 2020.

[9] C. Wang, L. Liu, X. Hao, H. Guo, Q. Wang, J. Huang, N. He, H. Yu, X. Lin,A. Pan, et al., “Evolving epidemiology and impact of non-pharmaceutical interventions on the outbreak of coronavirus disease 2019 in wuhan, china,” medRxiv, 2020.

[10] M. G. Pedersen and M. Meneghini, “Quantifying undetected covid-19 cases and effects of containment measures in italy,” ResearchGate Preprint (online 21 March 2020) DOI, vol. 10, 2020.

[11] “Italian coronavirus cases likely “10 times higher than reported”,” Reuters, Mar. 2020.

[12] E. O. Nsoesie, R. Beckman, M. Marathe, and B. Lewis, “Prediction of an epidemic curve: A supervised classification approach,” Statistical communications in infectious diseases, vol. 3, no. 1, 2011.

[13] J.-P. Chretien, D. George, J. Shaman, R. A. Chitale, and F. E. McKenzie, “Influenza forecasting in human populations: a scoping review,” PloS one, vol. 9, no. 4, 2014.

[14] L. Magri and N. A. K. Doan, “First-principles machine learning modelling of covid-19,” arXiv preprint 2004.09478, 2020.

[15] R. Adhikari, A. Bolitho, F. Caballero, M. E. Cates, J. Dolezal, T. Ekeh, J. Guioth, R. L. Jack, J. Kappler, L. Kikuchi, H. Kobayashi, Y. I. Li, J. D. Peterson, P. Pietzonka, B. Remez, P. B. Rohrbach, R. Singh, and G. Turk, “Inference, prediction and optimization of non-pharmaceutical interventions using compartment models: the pyross library,” 2020.

[16] S. Zhao and H. Chen, “Modeling the epidemic dynamics and control of covid-19 outbreak in china,” Quantitative Biology, pp. 1–9, 2020.

[17] L. López and X. Rodo, “A modified seir model to predict the covid-19 outbreak in spain and italy: simulating control scenarios and multi-scale epidemics,” Available at SSRN 3576802, 2020.

[18] H. H. Ayoub, H. Chemaitelly, G. R. Mumtaz, S. Seedat, S. F. Awad, M. Makhoul, and L. J. Abu-Raddad, “Characterizing key attributes of the epidemiology of covid-19 in china: Model-based estimations,” medRxiv, 2020.

[19] X. Zhang, R. Ma, and L. Wang, “Predicting turning point, duration and attack rate of covid-19 outbreaks in major western countries,” Chaos, Solitons & Fractals, p. 109829, 2020.

[20] I. Ciufolini and A. Paolozzi, “Mathematical prediction of the time evolution of the covid-19 pandemic in italy by a gauss error function and monte carlo simulations,” The European Physical Journal Plus, vol. 135, no. 4, p. 355, 2020.

[21] R. Raziperchikolaei and H. Bhat, “A block coordinate descent proximal method for simultaneous filtering and parameter estimation,” in International Conference on Machine Learning, pp. 5380–5388, 2019.

